# Beyond Stigma: Family, Culture, and Mental Health Help-Seeking Among South Asian American Young Adults

**DOI:** 10.64898/2026.09.05.26362357

**Authors:** Sophia Khan, Xuewei Chen

**Affiliations:** Sam Houston State University College of Osteopathic Medicine, Conroe, Texas, USA; Department of Biomedical & Digital Health Intelligence in Clinical Systems, Sam Houston State University College of Osteopathic Medicine, Conroe, Texas, USA

**Keywords:** South Asian American, mental health stigma, help-seeking, culturally responsive care, family expectations

## Abstract

South Asian Americans are often aggregated within broader Asian American research on mental health help-seeking, potentially obscuring culturally specific family and community processes. This study examined how family systems, sociocultural expectations, and family roles shape recognition of emotional distress and pathways to mental health care among South Asian American young adults.

Five semi-structured virtual focus groups were conducted with 23 South Asian American young adults (ages 18-30) residing in Texas. Discussions explored mental health stigma, family influences, help-seeking, barriers to care, and strategies for improving mental health support. Transcripts were analyzed using MAXQDA, with analytic memos and reflexive practices informing theme development.

Nine interconnected themes emerged. Participants described community stigma becoming internalized through expectations of self-reliance, perfectionism, and family responsibility, shaping whether distress was recognized as warranting professional support. Families functioned as both barriers and facilitators of help-seeking, while intergenerational differences and mental health literacy influenced recognition and treatment acceptance. Older siblings described greater caregiving responsibilities and serving as early mental health advocates, while younger siblings described benefiting from increased family openness established by older siblings. Participants also emphasized privacy, culturally responsive care with cultural humility, and peer support through shared lived experiences.

These findings extend broader Asian American help-seeking literature by identifying how community stigma may become internalized through family and sociocultural expectations and highlighting sibling roles in shaping mental health attitudes within South Asian American families. Findings highlight culturally responsive care, family-centered mental health education, mental health literacy, and professionally facilitated peer support as potential pathways to care.

**What is the public significance of this article?:** This study highlights how family expectations, community stigma, and cultural norms can shape how South Asian American young adults understand mental health and seek support. The findings show that culturally responsive care, family-centered mental health education, and peer support groups could serve as potential strategies for improving mental health help-seeking and psychological well-being in South Asian American communities.

## Introduction

Mental health awareness and public discussion have increased among young adults in the United States (Saha, 2021), yet mental health service utilization remains disproportionately low among South Asians (Karasz et al., 2019). South Asian Americans also remain underrepresented in qualitative mental health research, limiting understanding of how cultural stigma, family expectations, and community influences are experienced and internalized in everyday life (Goel et al., 2023; Nhpang et al., 2025). Particularly little is known about how family roles such as birth order shape mental health attitudes and help-seeking or what culturally relevant strategies South Asian American young adults identify as potential pathways to care.

Previous research suggests that South Asian families and communities may place a strong emphasis on family honor, reputation, and collectivist values, which may contribute to mental health stigma and discourage professional help-seeking (Dalvi, 2024). These cultural and social dynamics may create unique barriers for South Asian American young adults in the United States navigating both traditional family expectations and Western social environments (Ball & Baidawi, 2026). Despite the rapid growth of South Asian American populations, this marginalized group remains largely underrepresented in mental health research (Nadimpalli et al., 2016).

Furthermore, South Asian Americans are often aggregated into broader Asian American categories in the current literature (Gordon et al., 2019). As a result, the considerable cultural, religious, linguistic, and family diversity within South Asian American communities is often understudied in literature, which may limit the development of culturally responsive mental health interventions, and produce a barrier to help-seeking (Xavier et al., 2026). Additionally, community and family stigma surrounding mental health concerns may discourage both open discussion and research participation, further limiting understanding of the mental health needs of this population (Habeb et al., 2025)

Therefore, this study sought to explore the lived, individual experiences of South Asian American young adults regarding mental health stigma, family expectations, birth order, help-seeking, barriers to care, and participant-identified strategies to improve mental health access. Using qualitative focus groups, this study aimed to better understand how family systems, cultural expectations, and community influences shape attitudes toward mental health and opportunities for culturally responsive interventions.

## Method

### Study design

This qualitative study employed five semi-structured virtual focus groups to explore mental health stigma, family dynamics, help-seeking, and participant-identified strategies to improve mental health care among South Asian American young adults. Participants were recruited from a previously completed mixed-methods survey examining mental health attitudes and barriers to care.

A qualitative approach was selected to better understand participants’ lived experiences and explore how family systems, cultural expectations, and community environments shaped mental health attitudes, barriers to help-seeking, and perceptions of mental health across generations. A semi-structured interview guide was developed to facilitate discussion while allowing participants to elaborate on topics of importance. Discussion domains included mental health discussions within families and communities, experiences of stigma, family expectations, family roles, birth order, help-seeking, cultural, financial, and structural barriers to care, navigating South Asian and Western cultural environments, culturally responsive mental health care, and participant-generated recommendations to improve mental health access. Follow-up questions and probing prompts were used throughout each focus group to clarify responses and explore emerging themes in greater depth.

### Participants and Recruitment

Participants were recruited from a previously completed mixed-methods study examining mental health attitudes and barriers to help-seeking among South Asian American young adults (ages 18-30 years) residing in Texas. Eligibility for the parent study required participants to self-identify as South Asian, be between 18 and 30 years of age, and reside in Texas. Recruitment for the parent study occurred through convenience sampling using community-based events, South Asian community organizations, university student organizations, and social media platforms throughout Texas, with a particular emphasis on communities within the Dallas-Fort Worth metropolitan area.

At the conclusion of the anonymous survey, participants were invited to indicate interest in participating in an optional virtual focus group. Of 227 survey responses, 191 participants completed the parent survey, and 51 respondents expressed interest in the optional focus groups and were subsequently contacted to schedule participation. Twenty-three participants ultimately enrolled and participated across five virtual focus groups (Figure 1). Following analysis of the five focus groups, recurring patterns were identified across participant discussions while also allowing attention to different experiences.

**Figure 1.**
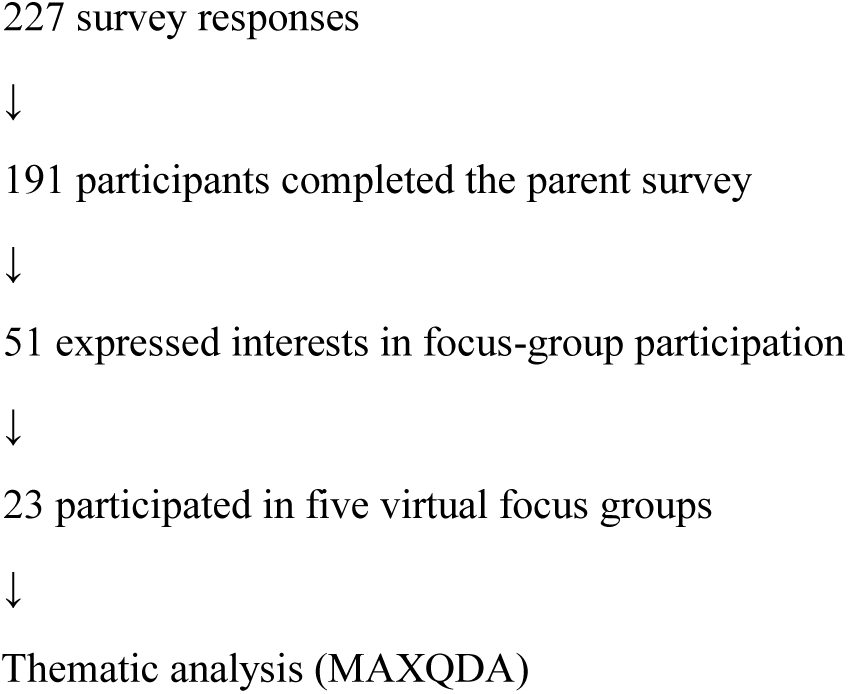
Participant flow diagram illustrating participant recruitment from survey completion through focus-group participation and thematic analysis.

### Focus groups

Five semi-structured virtual focus groups were conducted between June 10th and June 20th, 2026, with each session lasting approximately 60 minutes. Focus groups included three to seven participants to encourage discussion while allowing each participant to have adequate opportunities to share their experiences. All focus groups were conducted via Zoom by the primary researcher using a semi-structured interview guide, and the same moderator facilitated all five focus groups. Follow-up questions and probing prompts were used throughout each discussion to clarify participant responses and explore emerging topics in greater depth. This study is reported in accordance with the Consolidated Criteria for Reporting Qualitative Research (COREQ) guidelines. Before each focus group, participants were assigned de-identified participant numbers that were used through the discussion and transcription process to protect confidentiality. Audio and video recordings were transcribed prior to analysis.

### Data analysis

Audio and video recordings from all five focus groups were transcribed using Zoom Cloud and reviewed for accuracy before analysis. Transcripts were imported into MAXQDA (VERBI Software, Berlin, Germany) for qualitative analysis. Data were analyzed using thematic analysis by Braun and Clarke (2006). Initial codes were developed through repeated review of the transcripts and were iteratively refined as analysis progressed. Related codes were organized into broader categories, and themes were developed by identifying patterns across participants and focus groups. Coding and theme development were conducted by the primary researcher. Analytic memos were maintained throughout the coding process to document emerging interpretations, relationships among themes, and reflexive observations. Themes and interpretations were discussed with the senior author through the analytic process to enhance analytic rigor and challenge emerging interpretations. Following independent qualitative coding and initial theme development by the first author, ChatGPT (OpenAI) was used as a supplementary analytic tool to review selected de-identified excerpts and associated codes and provide feedback on coherence of the preliminary thematic organization. This tool was not provided with participant identifiers and did not independently conduct the initial coding or determine the final themes. All AI-generated feedback was critically reviewed by the first author, who retained responsibility for all analytic decisions, interpretation of participant narratives, and final theme development.

### Reflexivity

The primary researcher is a South Asian American medical student with an interest in mental health disparities among South Asian communities. Because the researcher shared aspects of participants’ cultural background, reflexivity was practiced throughout data collection and analysis to acknowledge how prior experiences and assumptions could influence interpretations, distinguish participant perspectives from researcher assumptions, and support the iterative development of themes. Throughout the analytic process, themes, codes, and interpretations were discussed with the senior author, who did not share the participants’ cultural background, to encourage reflexive dialogue, consider alternative interpretations, and enhance analytic rigor.

### Ethics

This study was approved by the [masked for review] Institutional Review Board (IRB [masked for review]). All participants provided informed consent prior to participation. Survey responses were collected anonymously and contained no personally identifiable information. Participants who volunteered for the optional focus groups provided contact information solely for scheduling purposes. These identifiers were stored separately from survey responses and removed from study data before qualitative analysis to protect participant confidentiality. During transcription, participants were assigned de-identified participant numbers, and identifying information was removed from transcripts whenever possible. Participation was voluntary, and participants could decline to answer any question or withdraw from the study at any time without penalty.

## Results

### Theme 1: Community Stigma Remains a Central Barrier to Mental Health Care

Participants consistently described community stigma as one of the most significant barriers to acknowledging emotional distress and seeking mental health care. Mental health concerns were frequently perceived as signs of weakness, poor character, lack of resilience, or personal failure. Participants perceived fears of judgement, gossip, and negative social perceptions if they disclosed emotional distress or sought professional support. These concerns extended beyond immediate family members and reflected broader community norms surrounding mental health, strength, resilience, and perceptions of those who seek professional support.

Representative quote:

> “Weakness was seen as not being strong enough to handle issues on your own. If you brought it up, you were seen as weak and not capable of dealing with things yourself.”

Collectively, participants consistently reported community stigma as reinforcing expectations of strength, resilience, and self-sufficiency while discouraging open discussion of emotional distress. These narratives suggest that concerns regarding community reputation and social perception extended beyond individual families and influenced how participants understood mental health within the broader South Asian community.

### Theme 2: Internalized Expectations and Help-Seeking

#### Stigma operates before treatment-seeking even occurs

Participants emphasized stigma as operating before treatment-seeking even occurred. Rather than functioning solely as a barrier to treatment or therapy, stigma influenced whether emotional distress was recognized as a legitimate concern in the first place. Participants described messages surrounding mental health becoming internalized, leading many to perceive emotional distress as a sign of weakness or personal failure.

Representative quote:

> “It’s very internalized. If we speak up, we’re seen as weak, so it makes us want to speak out less. If we can’t handle this by ourselves, how is that going to make us look in front of our parents or our community? It doesn’t just apply to mental health; it extends to other problems too. People don’t want to reach out because it’s so internalized that they’re thinking about how they’ll look in front of others.”

#### Internalized expectations shaped family roles and personal identity

Across multiple focus groups, participants consistently reported internalizing expectations regarding what it meant to be a “good” son or daughter. Rather than attributing these expectations solely to parents or family members, several participants consistently reported holding themselves to exceptionally high standards. Expectations to remain resilient, care for family members, succeed academically or professionally, and avoid burdening others became self-imposed. These expectations were frequently described as self-imposed rather than explicitly enforced by family members, suggesting that cultural expectations become internalized over time.

Representative quotes:

> “Taking care of your parents is just an understood thing in our family. It puts a lot of pressure on me to be the son my parents wanted. There’s this layer where you don’t want to say anything because it feels like it shows weakness. When I first saw a therapist, I didn’t tell anybody for quite a while.”
>
> “For me, it’s less family expectations more me being a perfectionist. I put the expectation on myself that I have to take care of my parents, my grandparents, and my younger sibling. In our South Asian community, a daughter is supposed to take care of her parents, so I put a lot of pressure on myself to act a certain way.”

Participants also described internalized expectations surrounding achievement and success. Several discussed believing they should avoid failure, solve problems independently, and continue meeting responsibilities despite emotional distress. One participant emphasized that making mistakes and needing additional support are normal parts of life, yet described these experiences as being difficult to accept because of deeply internalized expectations to remain resilient and self-sufficient.

Representative quote:

> “I remember telling myself that failure means you still learn something. I have to remind myself that I’m smart. Just because I didn’t get it on the first try doesn’t mean it’s not worth it. I just analyze what went wrong and try again.”

### Theme 3: Intergenerational Differences Shape Mental Health Attitudes

Participants frequently described a divide between younger and older generations regarding mental health awareness, willingness to discuss emotional concerns, and treatment acceptance. Many participants believed younger generations were becoming more open to mental health discussions while participants perceived older generations as holding more stigmatizing attitudes toward mental health. Participants often attributed these differences to varying levels of mental health awareness, differences in cultural norms surrounding emotional expression, and reduced access to mental health education among older generations. These differences were described as influencing family communication surrounding mental health and participants’ comfort discussing emotional concerns with parents and older relatives.

Representative quotes:

> “In my generation, there’s not an immediate place of judgment. Mental health isn’t a conversation that warrants opinion—it’s just people’s experiences”
>
> “I know it’s okay to not feel okay, but when I talk to my parents about it, they’re like, ‘When I was younger, that didn’t exist.’ No—it did exist. You just ignored it.”

Collectively, these quotations suggest that intergenerational differences may influence how mental health stigma is transmitted or challenged within families. While younger generations described greater openness toward discussing mental health, participants often continued to navigate older family members’ beliefs, creating tension between evolving attitudes and longstanding cultural expectations.

### Theme 4: Family Systems Function as Both Barriers and Facilitators to Help-Seeking

Although many participants described family-related barriers, others described parents or family members becoming more understanding over time through mental health education, treatment exposure, or open discussion. These accounts suggest that family attitudes were not static and could evolve, allowing families to function as facilitators as well as barriers to help-seeking.

Representative quote:

> “I’ve had trouble talking to them about mental health, or they’ve dismissed me in ways where they’re like there’s no such thing. They basically would say, like, doesn’t exist. And so, trying to have that conversation with them has been difficult.”

Representative quote:

> “My family has always been, again, really supportive, and they’re always like, if you need to talk about some things and just tell us, and, and you know, we’ll help you in any way.”

Collectively, these findings suggest that family systems function as the primary context through which mental health beliefs are transmitted, reinforced, and, in some cases, transformed. As a result, families may either perpetuate stigma or facilitate help-seeking depending on their level of openness, mental health awareness, and willingness to engage in conversations surrounding emotional distress.

### Theme 5: Mental Health Literacy

Participants frequently discussed a lack of understanding regarding mental health conditions and when professional support should be sought. Many described uncertainties about what constitutes a mental health concern worthy of intervention.

Representative quote:

> “The South Asian community often is very reactive with healthcare, and mental health is a big part of that. We don’t address mental health until it becomes very severe or even tragic. Even when physical symptoms appear, we often don’t seek treatment until they become severe or irreversible.”

Many participants described a cultural tendency to prioritize physical health while minimizing mental health or emotional wellbeing. Depression, anxiety, and other mental health concerns were frequently described as being viewed as less legitimate than physical illnesses. Participants repeatedly emphasized the need to recognize mental health as an essential component of overall health in addition to physical health.

Families frequently met participants’ physical and material needs while emotional needs remained less recognized or prioritized. Participants described this as reflecting different cultural understandings of what constitutes legitimate healthcare rather than the absence of care or concern.

Representative quote:

> “Mental health should be treated the same way as physical health. If someone has a physical injury, they seek treatment. Mental health issues should be seen in the same light.”

#### Recognition of need differed from awareness of available services

Participants often appeared less concerned with knowing that therapy existed than with recognizing when emotional distress warranted professional support. Several responses suggested that emotional distress is frequently normalized or minimized, making it difficult to determine when professional support may be appropriate. Collectively, these findings suggest that mental health literacy extended beyond knowledge of available services and included recognizing emotional distress as a legitimate health concern worthy of professional support.

### Theme 6: Culturally Responsive, Patient-Centered Care

Participants frequently expressed the importance of culturally responsive mental health care and described cultural understanding as an important mechanism for establishing trust and facilitating open discussions about mental health. Many participants preferred South Asian or culturally concordant providers because they believed these clinicians would better understand community-specific values, family dynamics, religious beliefs, and stigma, reducing the need to explain culturally specific experiences.

However, participants did not universally believe that providers needed to share their ethnic or cultural background. While some specifically preferred South Asian therapists, others emphasized that openness, cultural humility, empathetic listening, and a willingness to understand their lived experiences were equally important.

Representative quotes:

> “One thing I hesitated on was being a minority because I didn’t feel like they would fully understand where I was coming from. I specifically tried to find a Muslim South Asian woman who could relate to my experiences and understand my struggles.”
>
> “Someone of South Asian descent may be more likely to understand those challenges, but I don’t think the provider’s background always matters. If they’re open, empathetic, and willing to understand, they don’t have to go through the same experiences. At the end of the day, it boils down to the individual.”

### Theme 7: Community Judgment, Family Image, and Privacy-Shaped Help-Seeking

Concerns regarding reputation, family image, and *log kya kenghe* (“what will people think”) were prevalent throughout the dataset. Participants described concerns regarding gossip, community judgment, and maintaining family reputation with closely connected South Asian communities.

#### Anticipated judgment may be as powerful as actual judgment

Many participants discussed fear of judgment rather than specific experiences of judgment. This suggests that perceived social consequences alone may discourage treatment-seeking even in the absence of direct discrimination or criticism.

Representative quote:

> “People are afraid that if what they’re going through gets out, they’ll be judged, gossiped about, or pitied. Even if people don’t judge you, you’re worried that’s how they’ll see you.”

Participants consistently described concerns regarding privacy, confidentiality, and visibility within closely connected South Asian communities. Suggestions included private options, ways to hide insurance billing from family, anonymous services, and interventions that reduce community exposure.

#### Privacy concerns were closely intertwined with stigma concerns

Participants often described privacy not as a convenience but as a necessity for overcoming anticipated judgment, with anonymous and discreet treatment options viewed as strategies for reducing anticipated social consequences associated with help-seeking.

Representative quote:

> “I did all my therapy appointments in my car because I couldn’t do them at home. I had to explain to my therapist that I was okay, I just couldn’t have the appointment in my house. When you live with your parents, it’s difficult to keep things private if someone is listening on the other side of the door.”

### Theme 8: Shared Lived Experience Fosters Connection and Reduces Isolation

Participants consistently stated the focus group setting as a valuable opportunity to discuss mental health with other South Asian American young adults who shared similar cultural experiences. Many explained that hearing others describe comparable family dynamics and mental health challenges reduced feelings of isolation and created a sense of validation. Participants frequently distinguished peer support from formal psychotherapy, describing it as a complementary source of connection rather than a replacement for professional mental health care.

Several participants suggested that peer support groups centered on shared lived experiences could provide a culturally meaningful way to normalize conversations surrounding mental health, reduce isolation, and foster connection through shared cultural understanding.

Representative quotes:

> “Most South Asian people our age have either gone through something similar or know someone who has. There’s a commonality between all of us, and I think that’s what makes our generation unique. Being able to openly talk about it, knowing we share that foundation, is a good thing.”
>
> “Since it’s a South Asian group, there’s this relief that you don’t have to explain certain family or social dynamics—they’re already understood. If you say a couple of words, someone immediately understands what you mean. That’s what’s nice about these spaces.”
>
> “It’s nice to be in a setting with less pressure than an appointment. Whatever you say is understood within a cultural context, so you don’t have to explain everything. It’s not a one-stop solution, but it’s really nice to have a peer support group where you can talk about mental health concerns.”

Collectively, participants described shared lived experiences as reducing the need to explain culturally specific family dynamics or social expectations. Rather than replacing psychotherapy, peer support groups appeared to foster validation, normalize mental health discussions, and create culturally meaningful spaces where participants felt immediately understood.

### Theme 9: Birth Order, Family Roles, and Mental Health Advocacy

Participants stated birth order as influencing not only individual mental health experiences but also the evolution of family attitudes toward mental health over time. Older siblings frequently described serving as the first family member to navigate conversations surrounding emotional distress and professional help-seeking while simultaneously managing greater expectations related to caregiving, achievement, and serving as role models. Several participants stated these responsibilities as contributing to parentification, emotional burden, and reluctance to seek support for themselves.

In contrast, younger siblings frequently described benefiting from these earlier experiences. Rather than initiating conversations surrounding mental health, many described entering family environments in which older siblings had already challenged stigma, introduced therapy, or advocated for greater openness. Participants perceived this as resulting in fewer expectations surrounding emotional resilience and greater acceptance of mental health discussions within the family. Many younger siblings perceived themselves as benefiting from family changes initiated by older siblings, describing fewer expectations because older siblings had absorbed many of the responsibilities and challenges associated with introducing conversations surrounding mental health.

Participants who were only children described a distinct experience characterized by early independence, acting as the primary bridge between their parents and their own generation, and navigating family challenges without sibling support. Several described mediating family conflict independently while balancing expectations associated with being an only child.

Representative quotes:

> “I’m the youngest, so I feel like I got away with more. My older siblings had to navigate mental health without anyone to advocate for them. By the time it got to me, they were able to advocate for me. That’s why I think birth order definitely matters.”
>
> “In South Asian families, age hierarchy is a big thing. There are more expectations placed on older siblings, and it’s harder to talk about your own mental health struggles because you’re expected to be tough.”
>
> “Being the eldest daughter leads to chronic burnout and perfectionism. You’re essentially acting like a second mother even though you didn’t sign up for it. I grew up hyper-independent because I was expected to take care of everything on my own.”
>
> “I’m an only child, and because both of my parents were working professionals, I had to learn to manage everything on my own from an early age. That independence also helped my parents recognize that I was struggling and become more understanding.”

## Discussion

This qualitative study explored how help-seeking among South Asian American young adults is shaped by interconnected influences in which community expectations become internalized through family systems, influencing how participants recognized and responded to emotional distress (Figure 2). Participants reported mental health stigma as extending beyond treatment-seeking and becoming internalized through expectations surrounding strength, resilience, and self-reliance. Family systems, intergenerational differences, and birth order shaped how these expectations were experienced, while culturally responsive care and opportunities for shared lived experience emerge as potential pathways for improving engagement with mental health support.

**Figure 2.**
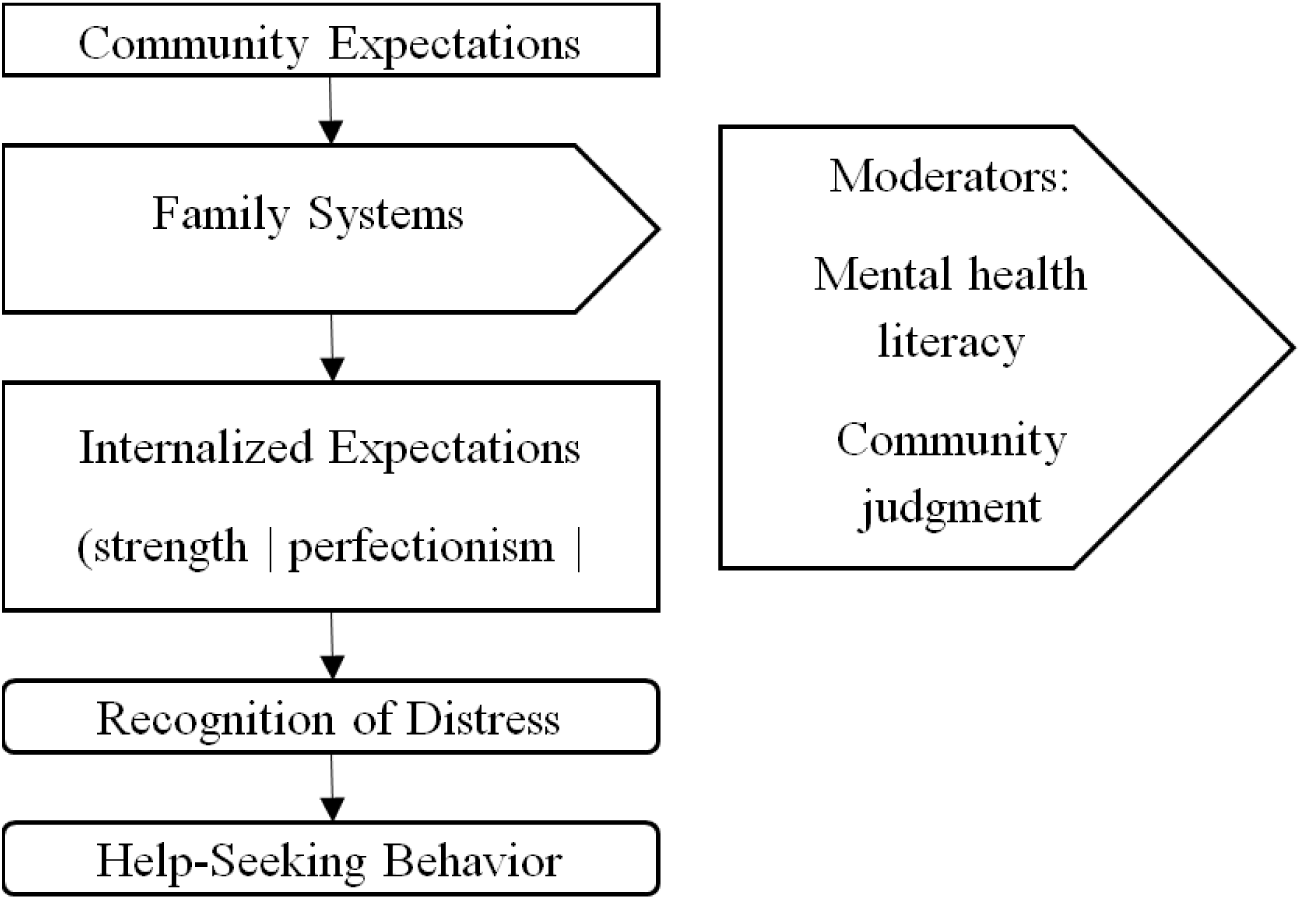
Conceptual model illustrating the relationships among community expectations, family systems, internalized expectations, recognition of emotional distress, and help-seeking among South Asian American young adults. Contextual influences identified through thematic analysis are shown alongside the primary conceptual pathway. ChatGPT (OpenAI, 2026) was used to assist with the visual organization and layout; all final content and design decisions were made by the authors.

Throughout the focus groups, participants consistently described mental health concerns and help-seeking as signs of weakness or an inability to manage problems independently. Rather than functioning solely as barriers to treatment, participants emphasized these beliefs becoming internalized over time, contributing to self-reliance, perfectionism, and reluctance to acknowledge emotional distress. These findings suggest that community stigma may influence help-seeking not only through external social pressures but also through the gradual internalization of expectations surrounding strength and resilience (Nguyen et al., 2024). Uppal et al. (2025) described this phenomenon through the concept of reflected shame, whereby individuals perceive mental health concerns as a threat to family reputation and social standing. Other studies have suggested that tensions between collectivist family values and individualistic cultural norms in the United States may further contribute to stigma surrounding emotional expression and help-seeking (Uppal et al., 2025). Additionally, internalization of the model minority myth has been associated with increased pressure to achieve, feelings of insecurity, and reduced utilization of mental health services (DeVitre & Gloria, 2025; Uppal et al., 2025). The findings of this study extend the literature by suggesting a potential pathway through which community stigma becomes internalized. Participants perceived community messages surrounding strength, resilience, and self-sufficiency as gradually becoming self-imposed expectations that shaped how they interpreted their own emotional distress. Rather than simply delaying treatment, these internalized expectations appeared to shape whether participants recognized their distress as legitimate or deserving professional support. Although previous studies have examined perfectionism among Asian American populations, these studies have primarily conceptualized perfectionism as arising from academic and familial pressures and have linked it to depressive symptoms (Suh et al., 2022; Yoon & Lau, 2008). In contrast, participants in the present study described perfectionism as intertwined with mental health stigma and internalized expectations surrounding self-reliance, suggesting that these constructs may be more closely connected than previously described. Similarly, previous literature has reported preferences for managing emotional concerns within the family rather than through professional services (Uppal et al., 2025). Participants in the present study extended this finding by describing self-reliance as an internalized expectation associated with being resilient, independent, and a “good” son or daughter. Thus, stigma, perfectionism, self-reliance, and family expectations may operate as interconnected processes through which community norms shape recognition of emotional distress and help-seeking.

Birth order and sibling dynamics emerged as important influences on mental health attitudes and help-seeking among participants. Older siblings frequently described parentification, greater expectations to serve as role models, and greater difficulty discussing mental health or accessing professional support, whereas younger siblings often described benefiting from increased family openness and advocacy established by their older siblings. Only children similarly described assuming independent caregiving and culture-brokering roles within their families. Previous literature has documented parentification among older siblings in collectivist cultures, with first-born children frequently assuming caregiving responsibilities because of filial piety and family obligation and has examined siblings in immigrant families through the concept of culture brokering (Cho & Lee, 2019; Hafford, 2010). However, these constructs have rarely been examined in relation to mental health stigma and help-seeking among South Asian American families. Rather than functioning solely as caregivers or cultural brokers, participants in the present study described older siblings as the first family members to navigate mental health stigma, advocate for treatment, and facilitate greater openness toward mental health for younger siblings. These findings suggest that sibling relationships may represent an overlooked mechanism through which South Asian family systems evolve, influencing not only the distribution of family responsibilities but also the transmission of mental health attitudes and access to care across generations (Figure 4).

Mental health literacy emerged as another important theme throughout this study. Participants frequently described uncertainty regarding when emotional distress could be managed independently versus when professional intervention was appropriate. Many also perceived that physical health was prioritized over mental health within South Asian families and communities, contributing to delayed recognition of emotional distress as a legitimate health concern. Collectively, these findings suggest that mental health literacy extends beyond awareness of available services and includes recognizing emotional distress as worthy of professional support. Previous studies demonstrate that limited mental health literacy among South Asian populations contributed to delayed care and perceived need for treatment (Mohsin et al., 2025). Tse and Halsam further reported that Asian Americans often conceptualize mental illness more narrowly than White Americans, with this narrower conceptualization partially mediating reduced help-seeking attitudes independent of stigma (Tse & Haslam, 2021). Consistent with the DSM-5-TR, which recognizes that cultural contexts influence where the boundary between normality and pathology is drawn, participants in the present study described emotional distress as frequently normalized until it became severe or manifested physically (American Psychiatric Association, 2022). Prior research has likewise shown that South Asian individuals commonly express psychological distress through somatic symptoms and that physical health is often viewed as more legitimate than emotional well-being (Behera et al., 2026; Shaligram et al., 2022; Uppal et al., 2025). Our findings extend this literature by suggesting that the primary barrier may not simply be inadequate knowledge of mental health services, but rather difficulty recognizing emotional distress as a valid category of health requiring intervention. Participants repeatedly described self-reliance as the default response to emotional distress, a finding that aligns with recent evidence identifying self-reliance as the most frequently reported barrier among Asian Americans with unmet mental health needs (Vu et al., 2026). Together, these findings suggest that interventions focused solely on reducing stigma may be insufficient if emotional distress itself is not recognized as needing care. Instead, improving mental health literacy within South Asian communities may require addressing the cultural frameworks through which emotional distress is understood and legitimized.

Culturally responsive care emerged as another important theme throughout this study. Although many participants expressed a preference for South Asian mental health providers, others emphasized that shared ethnicity alone was not sufficient. Instead, participants consistently described cultural understanding, humility, empathy, and a willingness to learn about their lived experiences as more important than provider ethnicity itself. These findings are consistent with previous literature demonstrating that culturally responsive mental health care improved accessibility, engagement, and treatment outcomes among South Asian populations. Qualitative studies have highlighted the importance of culturally safe services and clinicians who understand family dynamics and cultural values, while culturally adapted interventions have demonstrated improved clinical outcomes compared with standard approaches (Husain et al., 2024; Menon et al., 2025; Naeem et al., 2024). Our findings extend this literature by distinguishing cultural concordance from cultural responsiveness. Although some participants preferred South Asian clinicians because they anticipated greater cultural understanding, many emphasized that empathy, openness, and cultural humility over shared ethnicity alone. This study’s participants seemed to distinguish between provider factual knowledge of South Asian culture and their dispositional qualities of humility and empathy. Participants described avoiding or delaying care not because providers were from different backgrounds, but because they anticipated their experiences would not be understood or contextualized within their cultural and family environments. These findings align closely with APA’s emphasis on cultural humility as an ongoing process of openness, self-reflection, and curiosity rather than mastery of cultural knowledge (American Psychological Association, 2021). Together, culturally responsive training may benefit from emphasizing curiosity, empathy, and cultural humility.

Another important finding of this study was the value participants placed on shared lived experience as a complement to traditional mental health care. Participants consistently described the focus group setting as providing a unique opportunity to discuss mental health with others who shared similar cultural experiences or family dynamics. Some participants preferred peer support groups composed specifically of South Asian young adults because of their shared cultural experiences. Others emphasized that shared ethnicity was less important than participating in a group where members felt comfortable openly discussing mental health. Participants also distinguished peer support from psychotherapy, emphasizing that these groups should not function as venues for advice-giving or therapeutic treatment, but rather as structured spaces for sharing experiences, reducing isolation, and fostering connection under the guidance of a trained mental health professional. Previous literature has consistently demonstrated that peer support improves psychological well-being, including reductions in depression, anxiety, and loneliness and improvements in coping and self-esteem among young adults and minority populations (Chien et al., 2019; Collado et al., 2026; Richard et al., 2022). Emerging evidence among Asian American populations similarly suggests that narrative sharing and community-based interventions reduce mental health stigma and strengthen community connectedness (Rivera et al., 2019; Tran et al., 2026). In addition, qualitative research has demonstrated that focus groups themselves may foster feelings of validation, empowerment, and group cohesion among participants (Sim, 1998). Our findings extend this literature by illustrating that participants valued peer support because it provided opportunities to witness and validate one another’s lived experiences rather than receive advice or psychoeducation. Participants repeatedly described the therapeutic value of being understood by individuals who require little explanation of their cultural experiences, suggesting that one mechanism underlying peer support may be validation and shared understanding rather than problem solving. Furthermore, participants consistently framed peer support as complementary to, rather than a replacement for, professional mental health treatment, suggesting that psychotherapy may address clinical needs while culturally grounded peer support addresses isolation, belonging, and shared lived experience. Together, these findings support future investigation of professionally facilitated peer-support interventions as a culturally responsive complement to psychotherapy for South Asian American young adults.

### Implications for Asian American Mental Health Practice

Clinical implications of this study include incorporating routine mental health screening into primary care visits, strengthening culturally responsive training for mental health professionals, engaging family systems in mental health education, and developing professionally facilitated peer-support groups for South Asian American young adults. Participants repeatedly described primary care as a potentially accessible setting for discussing emotional distress, particularly for individuals who may not independently recognize that their symptoms warrant mental health care or who perceive specialty mental health services as difficult to access. Embedding mental health screening into routine annual wellness visits may therefore provide a potentially accessible and de-stigmatizing entry point for earlier recognition of emotional distress and timely referral to mental health services. Participants also emphasized that culturally responsive care extends beyond provider ethnicity and instead requires cultural humility, empathy, openness, and a willingness to understand participants’ lived experiences. These findings suggest that clinician training should prioritize interpersonal skills alongside cultural knowledge to foster trust and therapeutic alliance. Given the prominent role of family systems, birth order, and intergenerational dynamics identified throughout this study, culturally tailored educational initiatives that engage family members may further reduce stigma and facilitate earlier help-seeking. Finally, participants consistently described professionally facilitated peer-support groups as valuable complements to traditional mental health treatment because they reduced isolation through shared lived experiences while maintaining appropriate clinical boundaries. Collectively, these findings support a multi-level approach to improving mental health care for South Asian American young adults that integrates primary care, culturally responsive clinical practice, family-centered education, and community-based peer support (Figure 3). These participant-informed recommendations are consistent with evidence supporting integrated primary care, culturally responsive behavioral health services, and peer-support interventions among underserved populations (Blackmore et al., 2022; Richard et al., 2022; Siddaiah et al., 2026).

**Figure 3.**
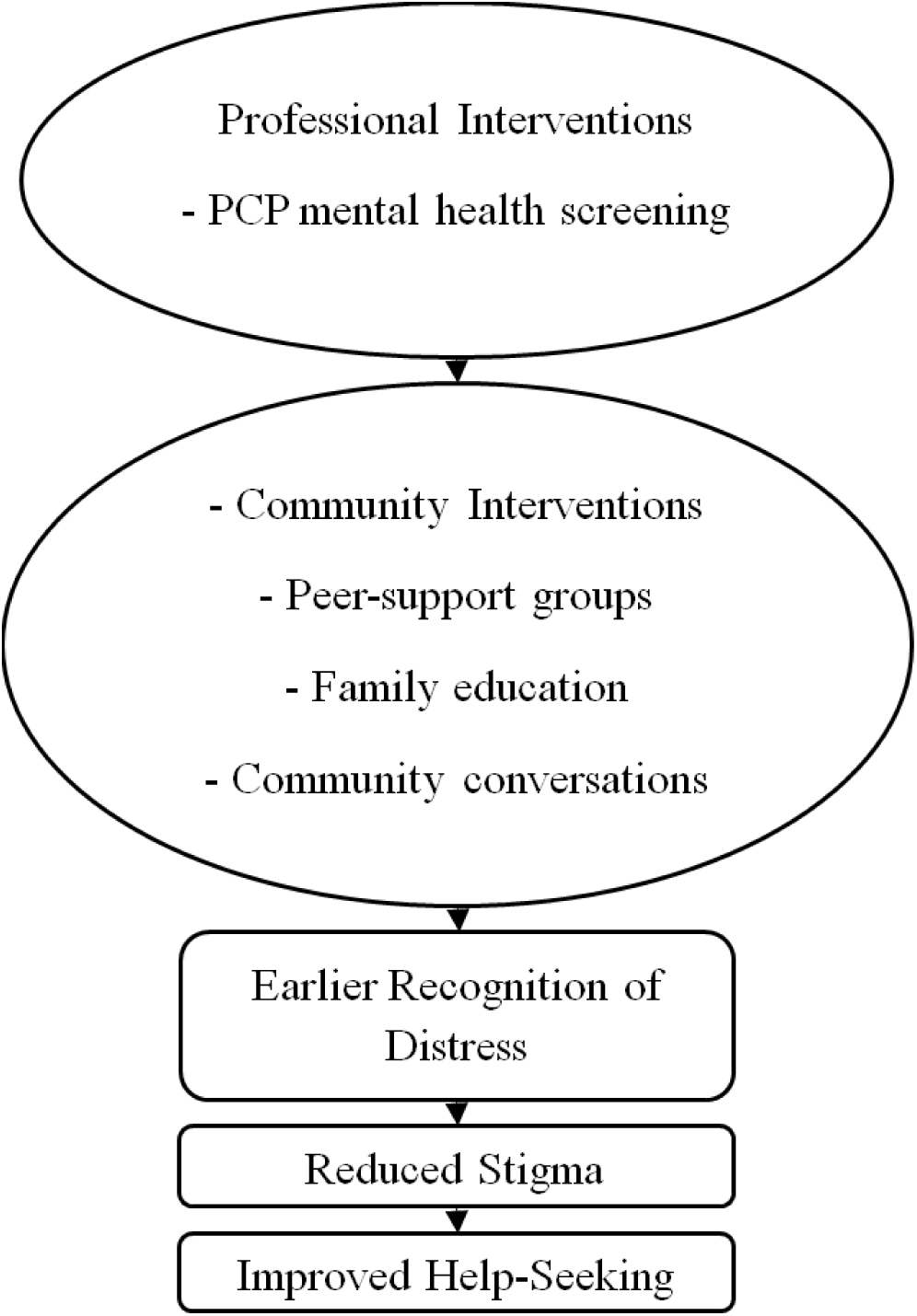
Participant-informed multilevel intervention framework summarizing participant recommendations for improving mental health support through clinical and community-based interventions. ChatGPT (OpenAI, 2026) was used to assist with the visual organization and layout; all final content and design decisions were made by the authors.

**Figure 4.**
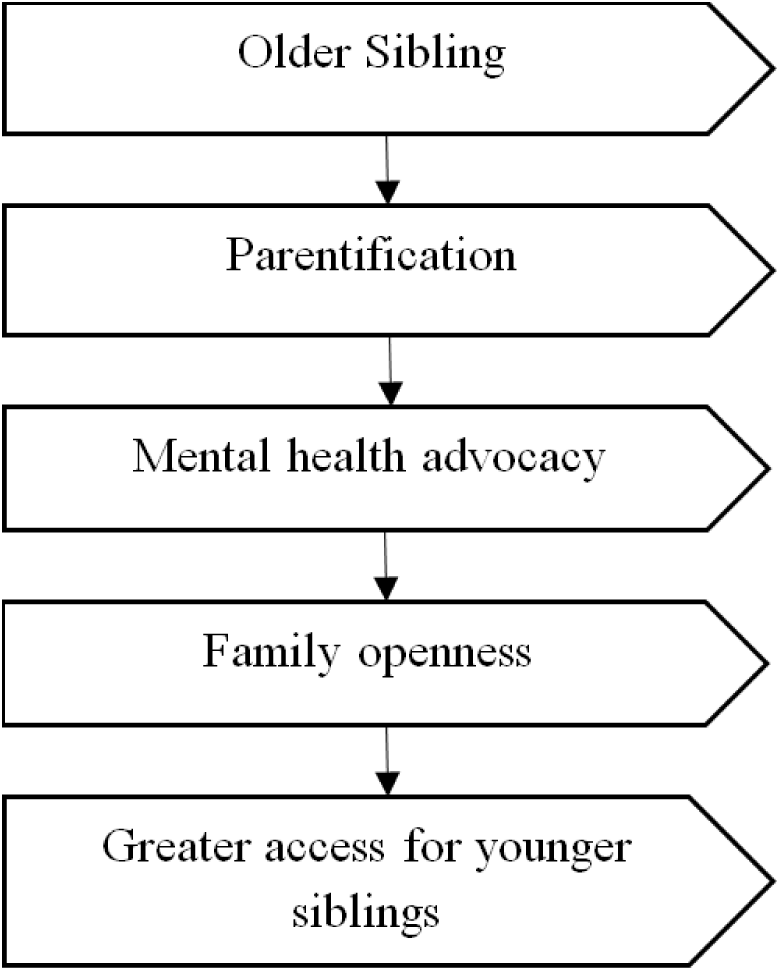
Family Systems and birth order model illustrating participants’ descriptions of older siblings serving as mental health advocates and facilitating greater family openness for younger siblings. ChatGPT (OpenAI, 2026) was used to assist with the visual organization and layout; all final content and design decisions were made by the authors.

### Strengths and Limitations

This study had several strengths, including its qualitative focus-group design, inclusion of an understudied South Asian American population, recruitment across community organizations, university networks, and social media rather than a single institution, and generation of participant-informed recommendations for future research and clinical practice.

This study had several limitations. First, participants were recruited exclusively from Texas therefore the findings may not be generalizable to South Asian communities in other regions of the United States, where cultural, religious, socioeconomic, and immigration experiences may differ. Additionally, South Asian American communities are heterogenous with respect to ethnicity, religion, language, immigration history, and socioeconomic background; the present sample may therefore not capture the full range of experiences within this population. Second, because participation in the focus groups was voluntary, individuals who chose to participate may have been more comfortable discussing mental health than those who declined, potentially limiting representation of individuals with the highest levels of stigma or reluctance to seek care. Third, the focus-group format may have influenced participant responses through group dynamics, social desirability, or conformity, despite efforts to foster an open and confidential environment through de-identification and skilled moderation. Furthermore, coding and theme development were conducted primarily by one researcher; however, themes and interpretations were discussed iteratively with the senior author to enhance analytic rigor. Participants may also have withheld particularly sensitive experiences because discussions occurred in a group setting rather than individual interviews. Finally, as with qualitative research, the findings are intended to provide an in-depth understanding of participants’ lived experiences rather than produce statistically generalizable estimates.

### Future Directions

Future research should examine whether these themes are replicated in larger and more geographically diverse South Asian American populations. Longitudinal and mixed-methods studies may further clarify how family systems, birth order, and internalized expectations influence mental health attitudes and help-seeking over time. Intervention studies should evaluate culturally responsive provider training, family-centered mental health education, primary-care screening, and professionally facilitated peer-support models. Similar qualitative approaches in other underserved populations may further identify both shared and culturally specific pathways influencing mental health care utilization.

## Conclusion

This qualitative study explored the lived experiences of South Asian American young adults and examined how family systems, cultural expectations, community norms, and personal experiences shape mental health attitudes and help-seeking. Participants reported mental health stigma as extending beyond barriers to treatment and becoming internalized through expectations surrounding strength, self-reliance, family responsibility, and resilience. These findings demonstrate that help-seeking is shaped by interconnected family, cultural, and community influences rather than individual attitudes alone. Participants also identified culturally responsive care and shared lived experiences as important facilitators of mental health support.

These findings suggest that improving mental health outcomes for South Asian American young adults will require interventions that extend beyond stigma reduction alone. Participants emphasized the importance of culturally responsive care grounded in cultural humility, routine mental health screening within primary care settings, family-centered mental health education, and professionally facilitated peer-support groups that complement traditional mental health treatment. Together, these findings highlight the importance of addressing both individual and family-level influences while creating culturally meaningful pathways to mental health care.

## Figures

Figure Shape Key

Rectangles = Risk factors/barriers

Rounded rectangles = Protective factors/outcomes

Oval = Intervention points

Pentagon = Contextual influence

## Data Availability

The datasets generated and/or analyzed during the current study are available from the corresponding author on reasonable request, subject to Institutional Review Board approval.

